# Mortality audit of cancer patients with SARS-CoV-2 positivity or COVID-19

**DOI:** 10.1101/2021.05.21.21257574

**Authors:** Kaberi Kakati, Tashnin Rahman, Debabrata Barman, Mouchumee Bhattacharyya, Bibhuti Bhusan Borthakur, Rashmisnata Barman, Apurba Kalita, Amal Chandra Kataki, Manigreeva Krishnatreya

**Author notes:** **Correspondence to**: Dr Manigreeva Krishnatreya, Room3, OPD Building, Dr. B Borooah Cancer Institute, Guwahati 781016, India, E-Mail.

## Abstract

Coronavirus disease-2019 (COVID-19) has disrupted cancer care services globally. The present review of cause of deaths was conducted in a tertiary care cancer center in the North East India. In our institute, all cancer patients requiring admission for surgery, chemotherapy, and other daycare procedures require testing for severe acute respiratory syndrome coronavirus-2 (SARS-CoV-2). From 09 July 2020 to 16 May 2021, 119 cancer patients with SARS-CoV-2 positive report or COVID-19 have been admitted at our institute Covid ward. A total of 19 cancer patients with COVID-19 succumbed. Of 19 deaths, 13 (68.4%) patients were men and 6 (31.6%) patients were women. The age range from 27 years to 74 years (median =55 years). Vomiting alone or with diarrhea was the most common symptom requiring admission after testing (4/19, 21.0%), followed by bleeding from primary tumour site (3/19, 15.7%). The antecedent and underlying cause of deaths in 19 (100%) patients was cancer. SARS-CoV-2 infection should not be a hindrance for cancer treatment and management.

## Introduction

The outbreak of second wave of severe acute respiratory syndrome coronavirus - 2 or SARS-CoV-2 has wreaked havoc in the country. The deaths due to coronavirus disease-2019 (COVID-19) have crossed 270,000 since the pandemic started in March 2020 in India [1]. According to the World Health Organization (WHO), death due to COVID-19 is defined for surveillance purposes as, “a death resulting from a clinically compatible illness, in a probable or confirmed COVID-19 case, unless there is a clear alternative cause of death that cannot be related to COVID disease (e.g. trauma)” [2]. In case of death due to COVID-19, there must not be a period of complete recovery from COVID-19 illness and death. A study has shown that acute respiratory failure and sepsis were the main causes of death in 77 cases based on patients’ clinical characteristics and laboratory results [3]. The reason for deaths in COVID-19 is suspected to be the “cytokine storm” or the systemic inflammatory response syndrome (SIRS) [4]. Excessive production of pro-inflammatory cytokines leads to acute respiratory distress Syndrome aggravation and widespread tissue damage resulting in multi-organ failure and death [5]. Early on during the pandemic, it was thought that cancer patients were susceptible to die due to COVID-19. However, our experience of last one year has shown that this is not true, and cancer treatment should not be halted in anticipation of cancer patients acquiring SARS-CoV-2 infection, except where it may be modified appropriately. We conducted a review of cause of deaths in admitted cancer patients with positive real time polymerase chain reaction (RT-PCR) or rapid antigen test (RAT).

## Materials and methods

This study has been done with approval from the Institutional Ethics Committee along with consent waiver. The present review of cause of deaths was conducted in a tertiary care cancer center in the North East India. In our institute, all cancer patients requiring admission for surgery, chemotherapy, and other daycare procedures require testing for SARS-CoV-2. A molecular virology laboratory was established for the same in July 2020. In case the person is tested positive for SARS-CoV-2, he or she is admitted in the COVID ward, which has 37 beds. From 09.07.2020 to 16.05.2021, 119 cancer patients with SARS-CoV-2 positive report or COVID-19 have been admitted at our institute’s Covid ward. Of them, 19 (15.9%) cancer patients with SARS-CoV-2 positive report or with COVID-19 have succumbed to their illness. In the first wave, from 09.07.2020 to 30.09.2020 (more than 2 months), ten cancer patients with COVID-19 have died. However, in the second wave, from 28.04.2021 to 16.05.2021, nine cancer patients with COVID-19 died in a span of 18 days. In the present review of death audit, we examined age, gender, main symptom that required admission, primary site of cancer, presence of co-morbidities, duration of death following admission, and presence of fever or influenza like illness. The causes of death statements were recorded according to the guidelines of the WHO into immediate causes of death, antecedent cause of death, and underlying cause, and further relevant conditions that may have contributed to fatal outcome. The death report of each of these 19 cases was forwarded to the chairperson of the state COVID-19 death audit board (Annexure I) formed by the local government, and reporting of all COVID-19 deaths were statutorily required to report to the audit board. The cause of death was finally certified by the death audit board. Patients are numbered according to deaths in the timeline of the study. Patient 1 was first cancer patient with COVID-19 who succumbed at our Covid ward.

## Results

Of 19 deaths, 13 (68.4%) patients were men and 6 (31.6%) patients were women. The age range varied from 27 years to 74 years (median =55 years). Vomiting alone or with diarrhea was the most common symptom requiring admission after testing (4/19, 21.0%), followed by bleeding from primary tumour site (3/19, 15.7%). Also, three patients were admitted for routine or emergency surgery. These three were considered as asymptomatic SARS-CoV-2 positive cancer patients. Three (15.7%) cancer patients were admitted for surgical procedures as seen in table 1. The antecedent and underlying cause of deaths in 19 (100%) patients was cancer. One (5.2%) patient had renal insufficiency as the contributing cause of death with serum creatinine at 3.2mg/dl [Table 2]. Three (15.7%) patients had history of fever (Patients 13, 14 and 16). Their fever lasted for 1-2 days only. There was no predilection for the primary site of cancer for patients requiring admission who subsequently succumbed [Table 3]. Four patients had other co-morbidities like hypertension, diabetes mellitus, chronic obstructive pulmonary disease, and interstitial lung disease as shown on table 3. Deaths occurred within one day following admission to upto 30 days following post-operative complications. The median duration of death following admission was 12 days.

**Table 1:** It shows the age, gender, and main presenting symptoms of cancer patients detected with COVID-19 or SARS-CoV-2 positive report

| Patient | Gender | Main Symptoms/reason for Hospital admission |
| --- | --- | --- |
| Patient 1 | M | Bleeding from primary tumour site |
| Patient 2 | M | Intractable vomiting |
| Patient 3 | F | Shortness of breath |
| Patient 4 | F | Bleeding from primary tumour site |
| Patient 5 | M | Vomiting |
| Patient 6 | F | Vomiting |
| Patient 7 | M | Severe abdominal pain |
| Patient 8 | M | Extreme weakness |
| Patient 9 | M | Bleeding from primary tumour site |
| Patient 10 | M | Breathing Difficulty |
| Patient 11 | M | Vomiting and diarrhea |
| Patient 12 | F | Admission for intercostal water seal Drainage |
| Patient 13 | M | Diarrhea |
| Patient 14 | M | Admission for Surgery |
| Patient 15 | F | Admission for Surgery |
| Patient 16 | M | Admission for Surgery |
| Patient 17 | M | Extreme weakness and weight loss |
| Patient 18 | F | Severe abdominal pain |
| Patient 19 | M | Absolute dysphagia |

**Table 2:** The table shows the cause of death

| Patient2 | Immediate Cause | Antecedent cause | Underlying Cause | Other significant conditions |
| --- | --- | --- | --- | --- |
| Patient 1 | Haemorrhagic shock | Cancer |  |  |
| Patient 2 | Dyselectrolytemia | Cancer |  |  |
| Patient 3 | Respiratory insufficiency | Cancer |  |  |
| Patient 4 | Anemia | Cancer |  |  |
| Patient 5 | Gastric outlet obstruction | Cancer |  |  |
| Patient 6 | Cancer cachexia | Cancer |  |  |
| Patient 7 | Hepatic insufficiency | Cancer |  |  |
| Patient 8 | Neutropenia sepsis | Cancer |  |  |
| Patient 9 | Dyselectrolytemia | Cancer |  |  |
| Patient 10 | Respiratory insufficiency | Cancer |  |  |
| Patient 11 | Neutropenic enterocolitis | Cancer |  |  |
| Patient 12 | Respiratory insufficiency | Cancer |  |  |
| Patient 13 | Neutropenia sepsis | Muti-organ dysfunction | Cancer |  |
| Patient 14 | Sepsis | Cancer |  |  |
| Patient 15 | Respiratory insufficiency | Post-operative collapse of lung | Cancer |  |
| Patient 16 | Sepsis | Burst Abdomen | Cancer |  |
| Patient 17 | Cancer cachexia | Cancer |  |  |
| Patient 18 | Cancer cachexia | Cancer |  | Renal insufficiency |
| Patient 19 | Cancer cachexia | Cancer |  |  |

**Table 3:** It shows primary site of cancer and presence of co-morbidly (ies)

| Patient3 | Cancer site | Days from admission to death | Co-morbidity |
| --- | --- | --- | --- |
| Patient 1 | Oropharynx | 3 | None |
| Patient 2 | Gall bladder | 12 | None |
| Patient 3 | Supraglottis | 3 | *HTN |
| Patient 4 | Breast | 22 | None |
| Patient 5 | Stomach | 25 | None |
| Patient 6 | Rectum | 1 | None |
| Patient 7 | Ovary | 16 | None |
| Patient 8 | Myelofibrosis | 1 | None |
| Patient 9 | Tongue | 15 | None |
| Patient 10 | Lung | 3 | Interstitial Lung |

|  |  |  | Disease |
| --- | --- | --- | --- |
| Patient 11 | Testis | 9 | **COPD |
| Patient 12 | Lung | 30 | None |
| Patient 13 | Stomach | 1 | None |
| Patient 14 | Sigmoid<br>Colon | 19 | None |
| Patient 15 | Tongue | 18 | HTN+***DM |
| Patient 16 | Colon | 19 | None |
| Patient 17 | Hypopharynx | 3 | None |
| Patient 18 | Gall bladder | 5 | None |
| Patient 19 | Esophagus | 13 | None |

\*HTN=Hypertension, \*\*COPD=Chronic obstructive pulmonary disease, \*\*\*DM=Diabetes mellitus

## Discussion

This review of causes of death in cancer patients afflicted with COVID-19 is first of its kind. Though, apparently the proportion of fatalities (15%) looks higher than the case fatality of COVID-19 in general [6], but deep dive into the cause of death revealed that the case fatality is unlikely to be higher than general population. The present mortality audit revealed that cancer patient of any age can come with symptoms that require hospitalization and can be managed effectively by testing followed by admission in the Covid Ward. Such lateral integration of care for cancer patients is the need of the hour during the pandemic [7]. Significant proportion of SARS-CoV-2 infections is asymptomatic and in many cases it causes a milder form of disease (COVID-19) like fever, chills, cough and sore throat [8]. In our mortality review, cancer patients with unrelated symptoms with COVID-19 like bleeding from the primary cancer site and vomiting were predominant, and that lead to death. In the mortality review we observed that, 21% of deceased cancer patients with COVID-19 had co-morbidities. Hypertension was seen in 50% of patients with co-morbidities.

After reviewing history and clinical presentations in the present mortality review, it showed that cause of death of all patients admitted in our institute with COVID-19 was due to cancer. In the present mortality audit, the most common immediate cause of death was cancer cachexia (21%) followed by respiratory insufficiency in the absence of typical symptoms of COVID-19 pneumonia or ARDS. Though, there are opposing views regarding cancer cachexia as the cause of death and whether this is rather an epiphenomenon. However, it is postulated that increase in platelet number and activation pathways, and arrhythmias appear to be the cause of sudden cardiovascular events leading to death in cancer cachexia [9]. One of the parameters we looked at was history of fever. Typically, in deaths due to COVID-19 and SIRS, fever is seen to last for more than 7 days [10], and typical ground glass opacities involving bilateral lungs are seen. The short duration (maximum 2 days) of fever in our cohort of cancer patients with COVID-19 favored death due to underlying cancer and not due to COVID-19. Additionally, anecdotal evidences from larger centers treating patients with ARDS due to COVID-19 suggest severe body ache and weakness with long lasting fever (more than 7 days) as early signs of worsening of COVID-19 requiring ventilator support.

### Limitation of the study

The assessment of cause of death was purely based on history and clinical presentation, and serum pro-inflammatory biomarkers like ferritin, D-Dimer, troponin-I, pro-calcitonin, and IL-6 were not tested. We justify the lack of pro-inflammatory biomarkers testing in the absence of fever in over 84% of the patient cohort or fever lasting not more than 2 days in three patients.

## Conclusion

Catching SARS-CoV-2 infection while coming for cancer treatment and further management should not be viewed as devastating outcome for cancer patients. We should encourage cancer treatment despite an ongoing ravaging pandemic. This type of mortality audits help to quantify the actual mortality due to COVID-19 and shall guide in framing a sound public health policy to combat future viral pandemics, if any.

## Data Availability

Yes. In excel spreadsheet. Can be provided upon request

## Acknowledgement

Authors thank Jamil Ahmed Barbhuiya and Tarun Sonowal, medical social workers of the institute for helping families of deceased cancer patients with COVID-19 in their cremation or burial by following COVID death protocol mandated by the government.

